# Knowledge, attitudes, and practices related to ocular safety among maintenance workers in a Ghanaian university: A cross-sectional study

**DOI:** 10.64898/2026.09.01.26361906

**Authors:** Clavert Kwarteng, Florence Mawuli Brew, Emmanuel Owusu

## Abstract

Occupational ocular injuries are a preventable yet neglected public health problem, particularly in low- and middle-income countries. Maintenance workers are exposed to diverse ocular hazards daily, yet compliance with protective measures is consistently poor.

A descriptive cross-sectional study was conducted among 85 maintenance workers at the Maintenance and Essential Services Organization (MESO) of Kwame Nkrumah University of Science and Technology (KNUST), Ghana, recruited through stratified convenience sampling across seven occupational sections. A structured questionnaire assessed knowledge of ocular hazards and protective equipment, attitudes toward ocular safety, and safety practices. Data were analyzed using IBM SPSS version 26 (IBM Corp., Armonk, NY, USA); chi-square and Fisher’s exact tests assessed associations (p < 0.05). Participants were predominantly male (84/85, 98.8%), with a mean age of 44.5 ± 10.4 years. Overall knowledge was good (mean 9.40 ± 1.59 out of 11), but attitude and practice scores were average (2.78 ± 0.92 and 3.27 ± 0.93, respectively). Most workers correctly identified goggles and face shields as protective, but only about half recognized that ordinary sunglasses and spectacles offer inadequate protection. Although 97.6% (83/85) recognized the need for ocular protection, only 7.1% (6/85) reported consistent protective eyewear use, and fewer than half (45.9%, 39/85) had received formal ocular safety training. Routine general protective equipment use was significantly associated with ocular protection use (Fisher’s exact test, p = 0.011). Sand and dust particles were the leading causes of injury and only 25% (5/20) of injured workers sought formal care.

Workers demonstrated good knowledge but poor attitudes and practices toward ocular safety, suggesting that knowledge alone does not translate into protective behaviour even within a relatively well-resourced institutional setting. Findings suggest that limited access to task-appropriate protective eyewear may represent an important institutional barrier. Institutional PPE supply and section-specific safety training are essential to bridge this knowledge–practice gap.

## Introduction

Occupational hazards are any material, process, activity, or situation capable of causing accidents or disease in the workplace [1]. Globally, these hazards are major contributors to morbidity and economic loss, particularly in low- and middle-income countries (LMICs) where safety standards remain weakly enforced [2,3]. Worldwide, an estimated 2.78 million workers die annually from work-related accidents and diseases, and a further 374 million suffer non-fatal injuries [4]. Ocular injuries constitute a significant proportion of workplace injuries although they are largely preventable with appropriate protective measures [5].

Occupational ocular injuries comprise any injury to the eye and/or adnexa occurring in the workplace [6], many of which require medical intervention and may result in permanent vision loss. Combined with the loss of working hours, they impose a substantial economic and quality of life burden on the individual, their family, and the society [5]. Despite accounting for just about 0.27% of the total body weight, the eyes are the most commonly injured organ, second only to the hands and feet [7]. The adult workforce in LMICs is disproportionately exposed to occupational hazards [2,4], and evidence suggests that up to 90% of all occupational ocular injuries could be prevented with appropriate protective eyewear [5]. Therefore, it is important to investigate the knowledge, attitudes, and practices (KAP) of workers as they can provide insight into the extent of implementation and enforcement of laws and regulations governing occupational health and safety. Thus, the knowledge, attitudes and practices regarding ocular safety among those whose occupations predispose them to these risks can provide evidence of the need for further interventions such as education, legislation, policies and research which ensure safe working environments. Considering that the professions with highest susceptibility to ocular injury lesions include manufacturing, mining, plumbing electrical, welding and maintenance working activity [8], maintenance workers provide a unique opportunity for investigating ocular safety at the workplace.

While studies have examined ocular safety among welders, mechanics, and farmers in Ghana [9,10,11], little is known about ocular safety practices among university maintenance workers. University maintenance workers present a distinctive opportunity for occupational eye health research, because such maintenance units bring together several artisanal trades, electricians, welders, carpenters, mechanics and plumbers under one institutional roof allowing multiple hazard profiles to be studied within a single administrative setting. These workers are also typically more formally trained than their counterparts in informal and private sectors. In addition, their employment within a university may expose them more directly to the adoption and implementation of occupational health policy. Studying this group therefore tests whether KAP gaps persist even under relatively favourable structural conditions.

Understanding workers’ knowledge, attitudes, and practices toward ocular safety is essential for designing effective workplace interventions [3,12]. This study explored KAP related to ocular safety among maintenance workers at Kwame Nkrumah University of Science and Technology (KNUST), Ghana, with the aim of identifying gaps and informing institutional policy, practice and research on occupational eye health.

## Materials and Methods

### Study design and setting

A descriptive cross-sectional study was conducted at the Maintenance and Essential Services Organization (MESO), located within the KNUST campus in Kumasi, Ghana. The MESO manages building and vehicle maintenance, electrical works, plumbing, carpentry, painting, road maintenance, and air-conditioning and refrigeration services across the university.

### Study population and sampling

The study population comprised all 120 maintenance workers across seven operational sections: Electrical, Plumbing, Air-conditioning and Refrigeration, Roads and Culverts, Painting, Carpentry, and Masonry. The minimum sample size was calculated using the Cochran formula [13]: n₀ = Z²pq/e², with Z = 1.96, p = 0.50, q = 0.50, and e = 0.05, yielding n₀ = 384, corrected to 92 for the finite population. A proportionate stratified convenience sampling technique was employed. The total worker population was divided into seven strata corresponding to the operational sections, and the number of participants recruited from each stratum was proportional to its size relative to the total population (Table 1). Within each stratum, eligible workers present during the data collection period were recruited consecutively until the proportional quota for that section was met. This approach is consistent with established practice in occupational KAP studies conducted in settings where a complete sampling frame with random selection is not operationally feasible [14]. A final sample of 85 participants was achieved, yielding a response rate of 92.4%.

**Table 1.**
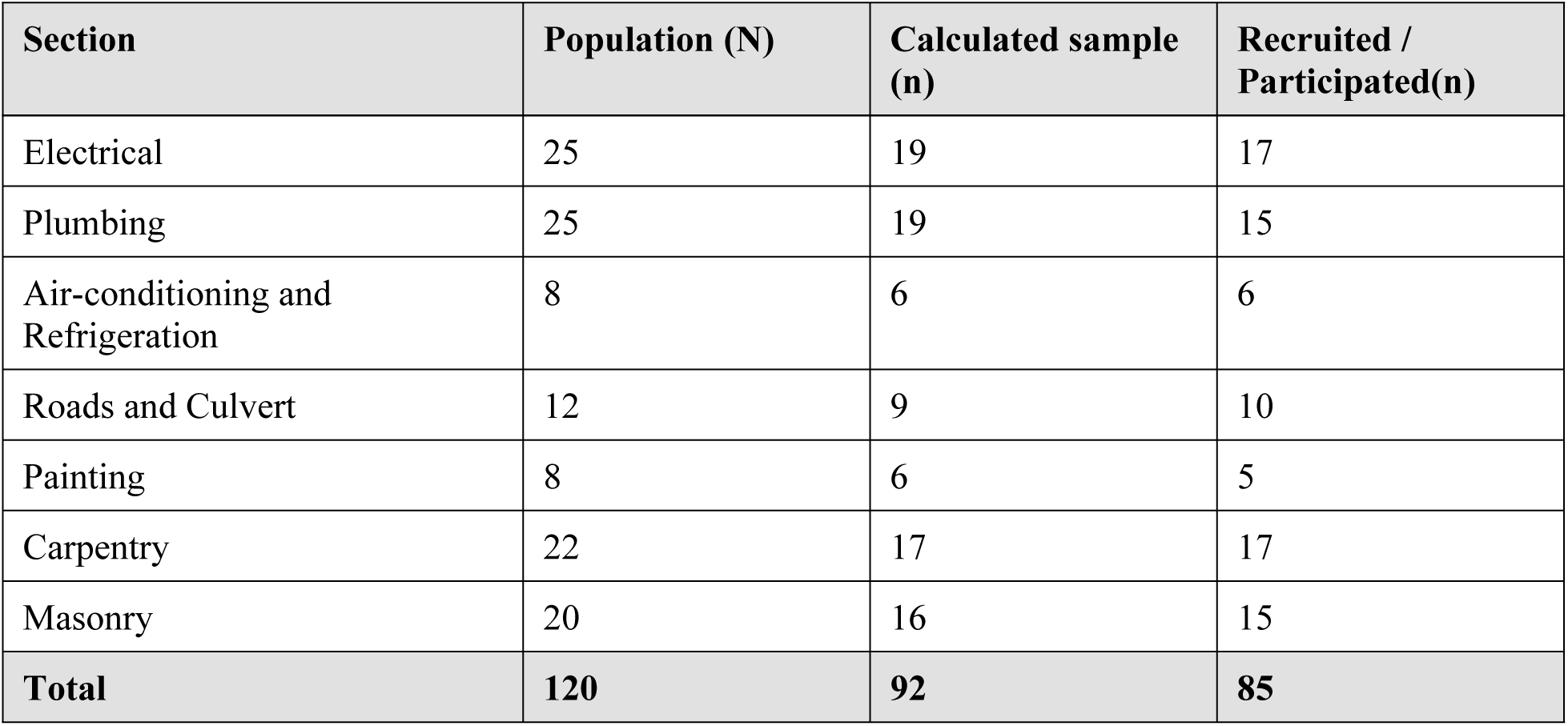
Distribution of maintenance workers across operational sections at MESO, KNUST, with calculated and actual sample sizes.

| Section | Population (N) | Calculated sample (n) | Recruited / Participated(n) |
| --- | --- | --- | --- |
| Electrical | 25 | 19 | 17 |
| Plumbing | 25 | 19 | 15 |
| Air-conditioning and Refrigeration | 8 | 6 | 6 |
| Roads and Culvert | 12 | 9 | 10 |
| Painting | 8 | 6 | 5 |
| Carpentry | 22 | 17 | 17 |
| Masonry | 20 | 16 | 15 |
| **Total** | **120** | **92** | **85** |

### Data collection

Data collection took place between 12 and 23 August 2021. Data were collected using a structured, pretested questionnaire administered via face-to-face interviews in English and the participant’s local dialect where necessary. All interviews were conducted by a single trained investigator, eliminating inter-rater variability. The questionnaire captured sociodemographic characteristics, knowledge of ocular hazards and protective equipment, attitudes toward ocular safety, and practices related to ocular protection. Information on occupational ocular injuries was also collected, including whether an injury had been experienced within the preceding 3 months, 6 months, or 1 year, along with the reported cause and management of the injury.

Knowledge, attitude, and practice responses were scored using a predefined scoring system. Each domain was categorized as poor, average, good, or excellent based on predetermined cut-off points specific to each scale as follows: knowledge of ocular hazards (5 items scored and graded as 0–1 = poor, 2–3 = average, 4–5 = good, 6 = excellent); knowledge of protective equipment (5 items scored and graded as 0–1 = poor, 2–3 = average, 4 = good, 5 = excellent); and total knowledge score (11 items scored and graded as 0–3 = poor, 4–6 = average, 7–9 = good, 10–11 = excellent). Attitude and practice scores were similarly categorized (0–1 = poor, 2–3 = average, 4 = good, 5 = excellent). These cut-off thresholds were adapted from scoring systems used in a prior KAP study on occupational eye safety in sub-Saharan Africa [11], and calibrated to the number of items in each subscale of the current instrument. The questionnaire was pretested among a similar group of workers who were not included in the final study to ensure clarity, relevance, and consistency.

### Ethical considerations

Ethical approval was obtained from the Committee on Human Research, Publication and Ethics (CHRPE) at the School of Medicine and Dentistry, KNUST (Reference number: CHRPE/AP/323/21). Written informed consent was obtained from each participant prior to data collection. Participation was voluntary, and confidentiality and anonymity were maintained throughout the study. The study was conducted in accordance with the principles of the Declaration of Helsinki.

### Data analysis

Data were entered into Microsoft Excel (Microsoft Corp., Redmond, WA, USA) and analyzed using IBM SPSS version 26 (IBM Corp., Armonk, NY, USA). Descriptive statistics (means, frequencies, percentages) were used to summarize participant responses. Chi-square tests were used for associations where all expected cell frequencies exceeded 5; Fisher’s exact test was applied where expected cell frequencies fell below 5, as recommended for small-cell contingency tables [15]. A p-value of less than 0.05 was considered statistically significant.

## Results

### Sociodemographic characteristics

Of the 92 eligible participants, 85 completed the questionnaire (response rate 92.4%). Eighty-four (98.8%) were male, and one (1.2%) was female. The mean age of the participants was 44.5 ± 10.4 years, ranging from 20 to 60 years. Carpentry and electrical sections each accounted for the largest occupational groups (20% each). The mean years of work experience was 15.0 ± 12.0 years. Over one-third (36.5%) of participants had tertiary-level education while only 3.5% had only primary education. The remainder had secondary level education.

### Knowledge, attitude, and practice scores

The mean knowledge, attitude, and practice scores, together with their corresponding interpretations, are summarized in Table 2.

**Table 2.** Mean knowledge, attitude, and practice scores and interpretations among maintenance workers at KNUST MESO.

| Domain | Mean Score (±SD) | Interpretation |
| --- | --- | --- |
| Knowledge of ocular hazards | 5.48 (±1.03) | Good |
| Knowledge of eye protective equipment | 3.92 (±1.03) | Average |
| Total knowledge score | 9.40 (±1.59) | Good |
| Attitude score | 2.78 (±0.92) | Average |
| Practice score | 3.27 (±0.93) | Average |

### Knowledge of ocular hazards and protective equipment

Workers demonstrated good knowledge of ocular hazards (mean 5.48 ± 1.03 out of 6). Dust (79/85, 92.9%), welder’s arc (84/85, 98.8%), and chemicals (75/85, 88.2%) were most widely recognized as ocular hazards (Fig 1). Knowledge of protective devices was moderate (mean 3.92 ± 1.03 out of 5): 81/85 (95.3%) correctly identified goggles and 77/85 (90.6%) identified face shields, but only 45/85 (52.9%) correctly recognized that non-prescribed sunglasses and spectacles offer inadequate occupational eye protection (Fig 2).

**Fig 1.**
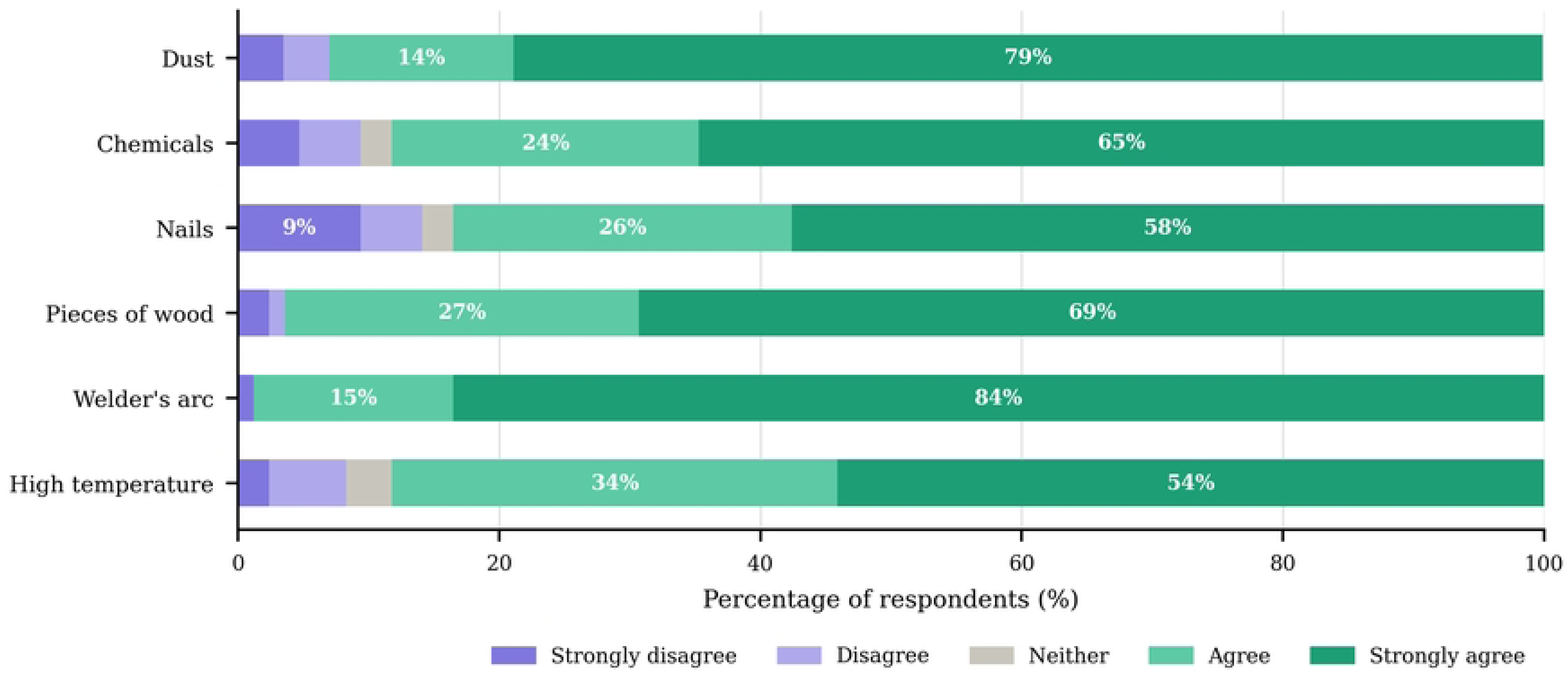
Knowledge of ocular hazards among maintenance workers at KNUST MESO (n = 85). Responses expressed as percentage of respondents per Likert category.

**Fig 2.**
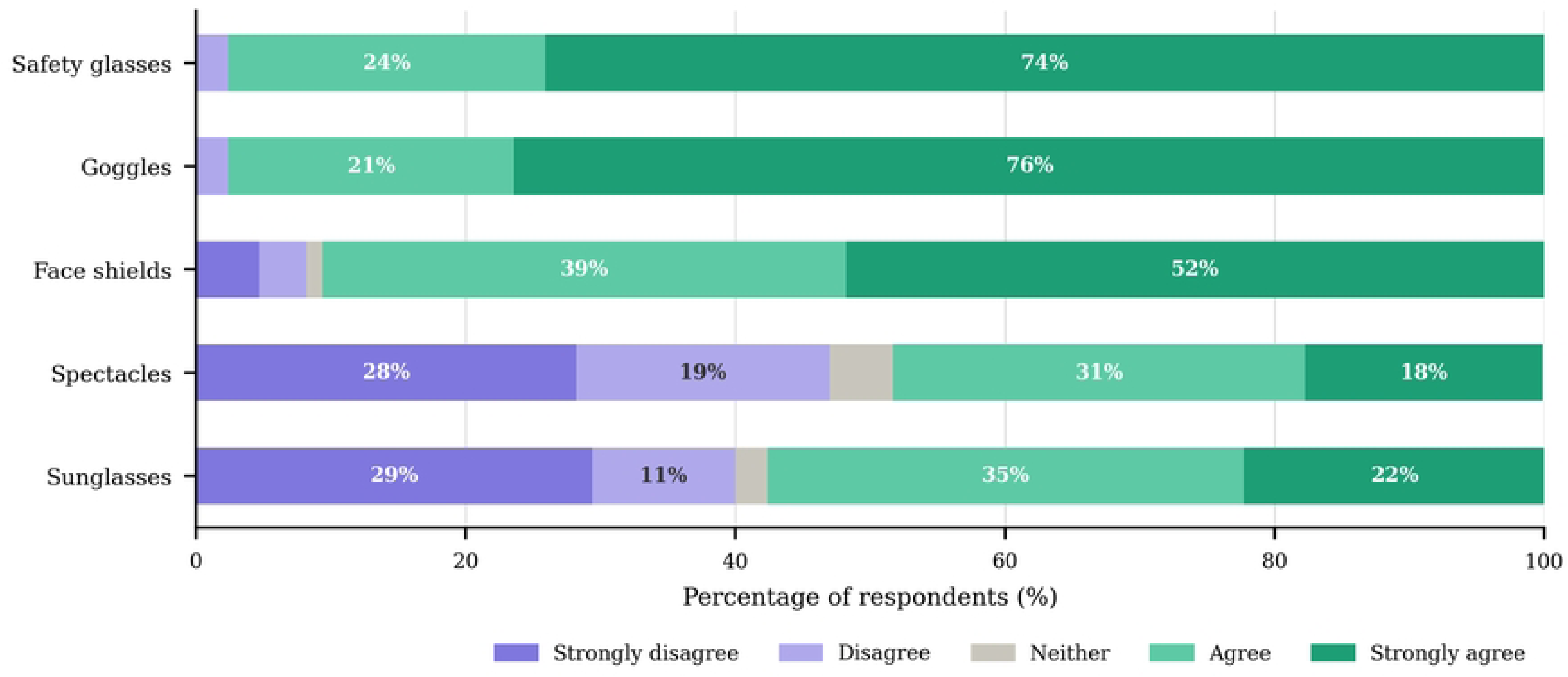
Knowledge of ocular protective equipment among maintenance workers at KNUST MESO (n = 85). Responses expressed as percentage of respondents per Likert category.

### Attitudes toward ocular safety

The vast majority (83/85, 97.6%) believed that protective eyewear is necessary; however, only 23.5% expressed willingness to personally purchase or contribute toward its procurement if necessary. Regarding enforcement of personal protective equipment use, most participants 98.0% considered penalties justified when workers refuse to use provided protective equipment (Fig 3).

**Fig 3.**
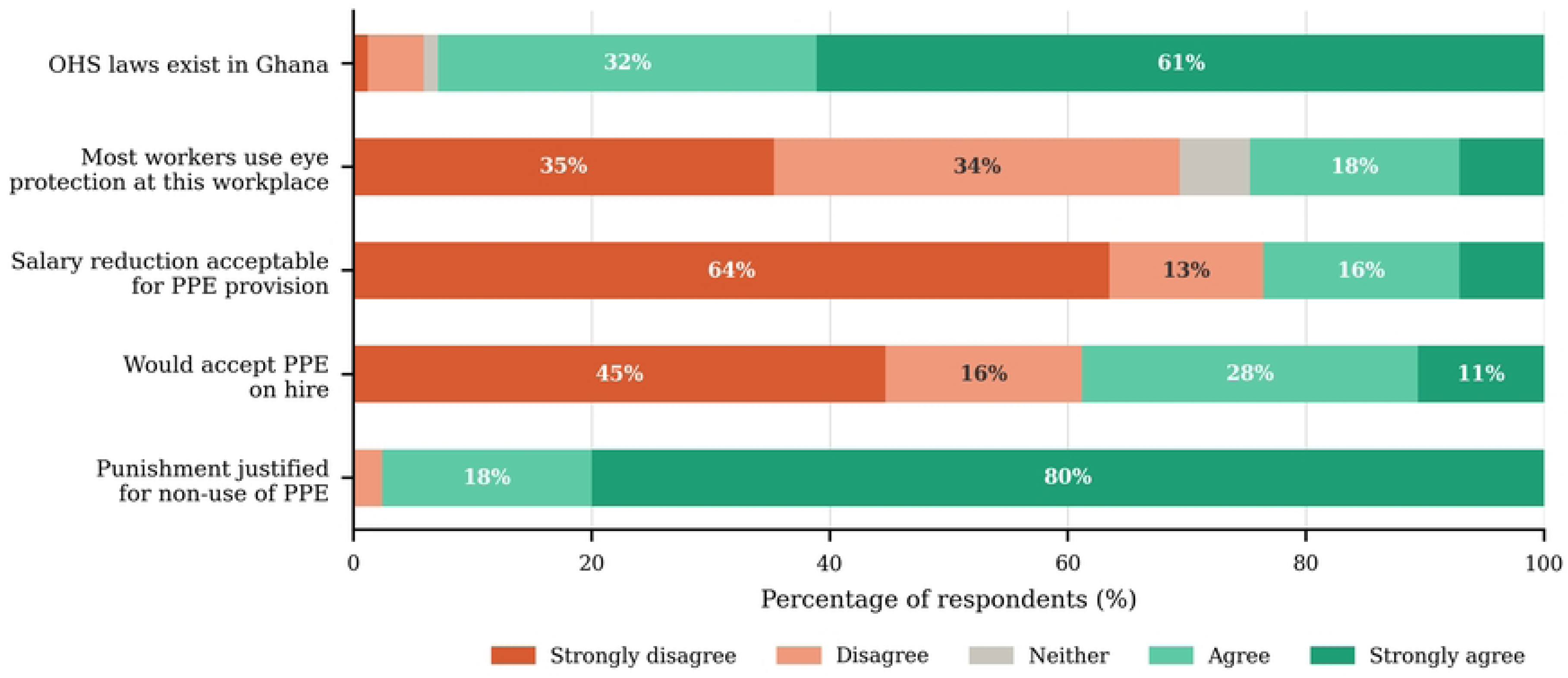
Attitudes of maintenance workers toward ocular safety at KNUST MESO (n = 85). Responses expressed as percentage of respondents per Likert category. OHS, occupational health and safety; PPE, personal protective equipment.

### Ocular safety practices

Only 39/85 (45.9%) of respondents had received formal ocular safety training. Despite 97.6% acknowledging the need for eye protection, only 14.1% reported having adequate protection for their specific work tasks, and only 7.1% reported consistent (always) use of protective eyewear (Fig 4). By contrast, 56.5% reported always using general protective gear. Most respondents (75.3%) obtained their protective equipment from MESO. Fisher’s exact test revealed a statistically significant association between routine use of general protective equipment and use of ocular protection (p = 0.011; Table 3). No significant associations were found between age (p = 0.372), occupation (p = 0.534), or educational level (p = 0.585) and the use of protective eyewear (Table 3). Among workers who used protective eyewear routinely, 40% were plumbers; no electrician or painter reported consistent eyewear use.

**Fig 4.**
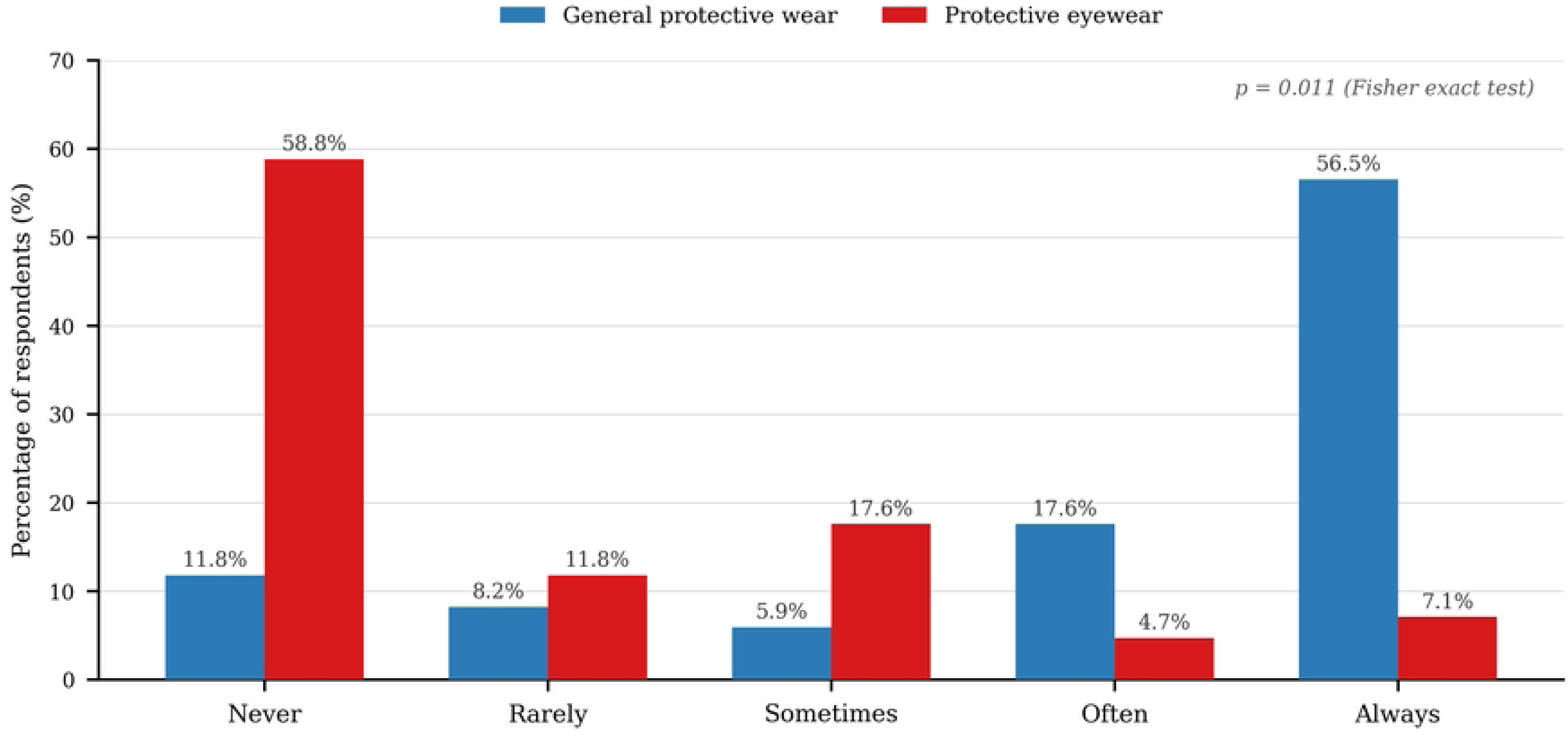
Frequency of general protective wear versus protective eyewear use among maintenance workers at KNUST MESO (n = 85). Fisher exact test: p = 0.011 for association between general PPE use and ocular protection use.

**Table 3.**
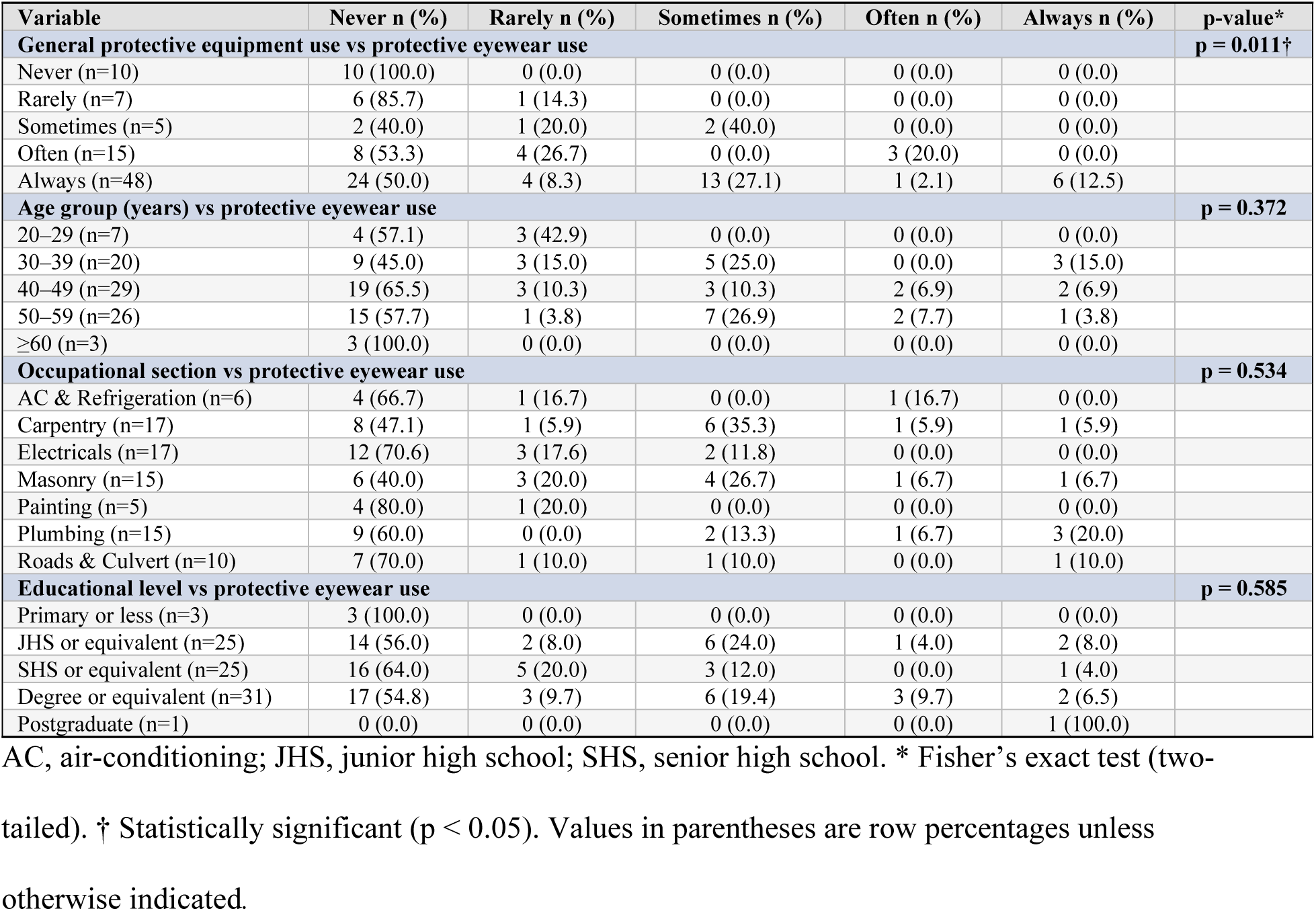
Associations between sociodemographic and occupational characteristics and protective eyewear use frequency, among maintenance workers at KNUST MESO (n = 85).

| Variable | Never n (%) | Rarely n (%) | Sometimes n (%) | Often n (%) | Always n (%) | p-value* |
| --- | --- | --- | --- | --- | --- | --- |
| **General protective equipment use vs protective eyewear use** |  |  |  |  |  | **p = 0.011†** |
| Never (n=10) | 10 (100.0) | 0 (0.0) | 0 (0.0) | 0 (0.0) | 0 (0.0) |  |
| Rarely (n=7) | 6 (85.7) | 1 (14.3) | 0 (0.0) | 0 (0.0) | 0 (0.0) |  |
| Sometimes (n=5) | 2 (40.0) | 1 (20.0) | 2 (40.0) | 0 (0.0) | 0 (0.0) |  |
| Often (n=15) | 8 (53.3) | 4 (26.7) | 0 (0.0) | 3 (20.0) | 0 (0.0) |  |
| Always (n=48) | 24 (50.0) | 4 (8.3) | 13 (27.1) | 1 (2.1) | 6 (12.5) |  |
| **Age group (years) vs protective eyewear use** |  |  |  |  |  | **p = 0.372** |
| 20–29 (n=7) | 4 (57.1) | 3 (42.9) | 0 (0.0) | 0 (0.0) | 0 (0.0) |  |
| 30–39 (n=20) | 9 (45.0) | 3 (15.0) | 5 (25.0) | 0 (0.0) | 3 (15.0) |  |
| 40–49 (n=29) | 19 (65.5) | 3 (10.3) | 3 (10.3) | 2 (6.9) | 2 (6.9) |  |
| 50–59 (n=26) | 15 (57.7) | 1 (3.8) | 7 (26.9) | 2 (7.7) | 1 (3.8) |  |
| ≥60 (n=3) | 3 (100.0) | 0 (0.0) | 0 (0.0) | 0 (0.0) | 0 (0.0) |  |
| **Occupational section vs protective eyewear use** |  |  |  |  |  | **p = 0.534** |
| AC & Refrigeration (n=6) | 4 (66.7) | 1 (16.7) | 0 (0.0) | 1 (16.7) | 0 (0.0) |  |
| Carpentry (n=17) | 8 (47.1) | 1 (5.9) | 6 (35.3) | 1 (5.9) | 1 (5.9) |  |
| Electricals (n=17) | 12 (70.6) | 3 (17.6) | 2 (11.8) | 0 (0.0) | 0 (0.0) |  |
| Masonry (n=15) | 6 (40.0) | 3 (20.0) | 4 (26.7) | 1 (6.7) | 1 (6.7) |  |
| Painting (n=5) | 4 (80.0) | 1 (20.0) | 0 (0.0) | 0 (0.0) | 0 (0.0) |  |
| Plumbing (n=15) | 9 (60.0) | 0 (0.0) | 2 (13.3) | 1 (6.7) | 3 (20.0) |  |
| Roads & Culvert (n=10) | 7 (70.0) | 1 (10.0) | 1 (10.0) | 0 (0.0) | 1 (10.0) |  |
| **Educational level vs protective eyewear use** |  |  |  |  |  | **p = 0.585** |
| Primary or less (n=3) | 3 (100.0) | 0 (0.0) | 0 (0.0) | 0 (0.0) | 0 (0.0) |  |
| JHS or equivalent (n=25) | 14 (56.0) | 2 (8.0) | 6 (24.0) | 1 (4.0) | 2 (8.0) |  |
| SHS or equivalent (n=25) | 16 (64.0) | 5 (20.0) | 3 (12.0) | 0 (0.0) | 1 (4.0) |  |
| Degree or equivalent (n=31) | 17 (54.8) | 3 (9.7) | 6 (19.4) | 3 (9.7) | 2 (6.5) |  |
| Postgraduate (n=1) | 0 (0.0) | 0 (0.0) | 0 (0.0) | 0 (0.0) | 1 (100.0) |  |

AC, air-conditioning; JHS, junior high school; SHS, senior high school. \* Fisher's exact test (two-tailed). † Statistically significant ( $p < 0.05$ ). Values in parentheses are row percentages unless otherwise indicated.

### Ocular injuries

A total of 20 respondents (23.5%) reported at least one ocular injury in the previous year. The leading causes were sand particles (30%), sawdust (15%), and dust (15%; Table 4). In terms of management, 60% irrigated the eye with water, while only three workers (15%) visited a hospital and two (10%) had visited a pharmacy (Table 5) for medication. For this study, formal eye care was defined as presentation to a hospital or pharmacy; self-treatment measures including eye irrigation, eye rubbing, and wiping with cloth were not considered formal eye care-seeking behaviours. Fisher’s exact test showed no significant association between occupation (p = 0.268), education (p = 0.657), or frequency of protective eyewear use (p = 0.147) and the occurrence of ocular injury. Given that only 20 injury events were recorded, the statistical power to detect such associations was limited, and these findings should be interpreted with caution.

**Table 4.** Causes of ocular injuries sustained in the past year among maintenance workers at KNUST MESO (n = 20).

| Cause of injury | n (%) |
| --- | --- |
| Sand particles | 6 (30.0) |
| Sawdust | 3 (15.0) |
| Dust | 3 (15.0) |
| Gas explosion | 2 (10.0) |
| Insect | 2 (10.0) |
| Chemicals | 1 (5.0) |
| Electric spark | 1 (5.0) |
| Glue | 1 (5.0) |
| Stone | 1 (5.0) |

**Table 5.**
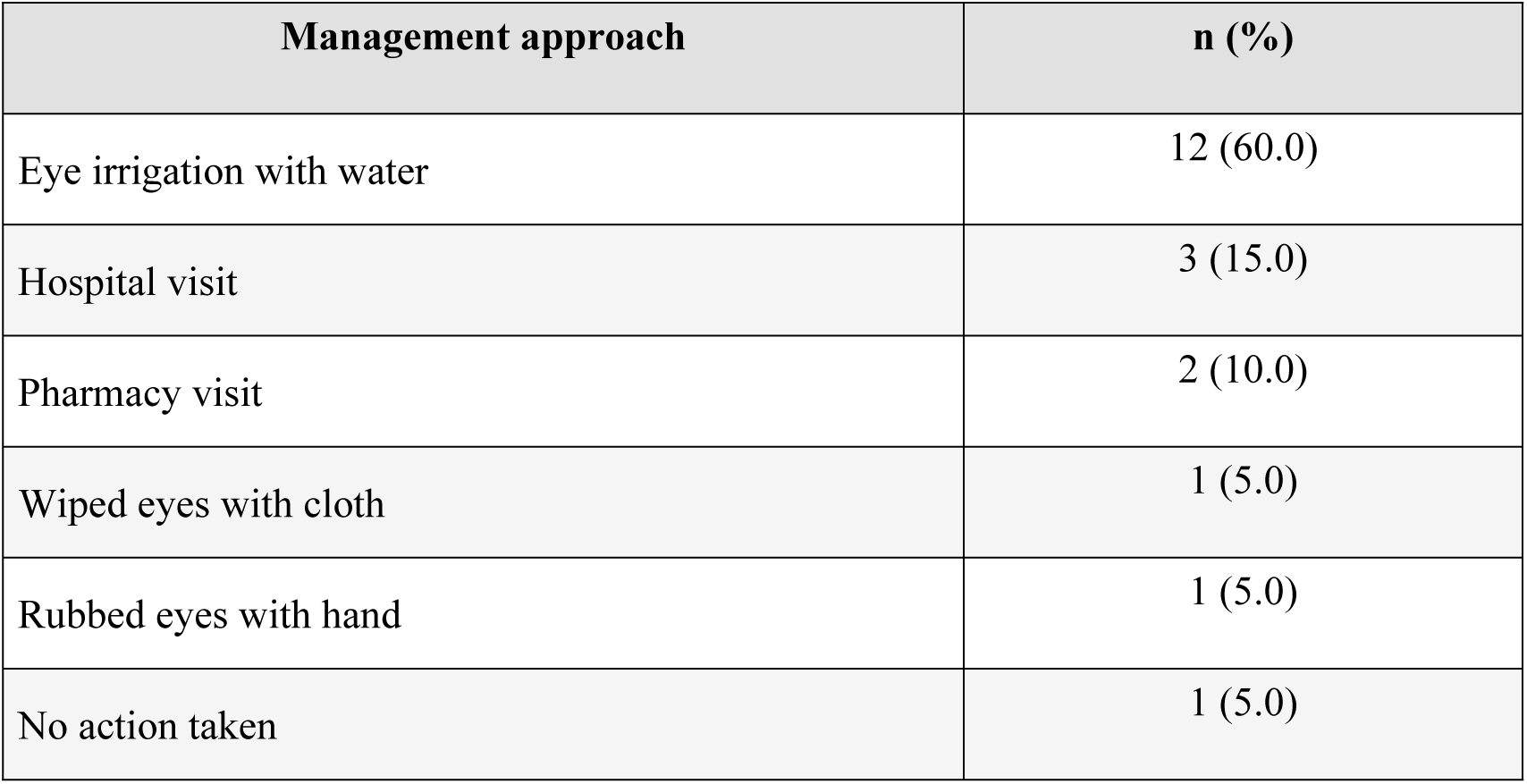
Management approaches taken following ocular injuries among maintenance workers at KNUST MESO (n = 20).

| Management approach | n (%) |
| --- | --- |
| Eye irrigation with water | 12 (60.0) |
| Hospital visit | 3 (15.0) |
| Pharmacy visit | 2 (10.0) |
| Wiped eyes with cloth | 1 (5.0) |
| Rubbed eyes with hand | 1 (5.0) |
| No action taken | 1 (5.0) |

## Discussion

Maintenance workers at the Kwame Nkrumah University of Science and Technology demonstrated good overall knowledge of ocular hazards and protective equipment (9.40 ±1.59 out of 11). This relatively high level of knowledge may be partly related to the educational and occupational characteristics of the study population. Most workers had attained at least secondary-level education, with over one-third (36.5%) holding a tertiary degree and only 3.5% having primary education or less. Higher educational attainment may facilitate understanding of workplace hazards and safety information, while employment within a university setting may also provide exposure to institutional health and safety policies that reinforce such knowledge. In addition, most workers had acquired their trade through apprenticeship, which may offer opportunities to learn about occupational hazards and safe work practices. Although these factors were not directly assessed in the present study, they may partly explain the relatively high knowledge scores observed, particularly in comparison with Ghanaian woodworkers, among whom the majority (77.3%) demonstrated low knowledge of occupational hazards [16].

Attitude scores were moderate overall (mean 2.78 ± 0.92 out of 5), although this overall score masked substantial variation across individual items. Nearly all workers (97.6%) agreed that protective eyewear is necessary, yet only 23.5% indicated a willingness to personally purchase or contribute toward its cost for their safety if they were not provided at the workplace. This contrast suggests that recognition of the importance of eye protection may not necessarily translate into willingness to invest into procurement of PPE. Similar patterns have been reported in previous studies, in which workers have generally regarded the provision of PPE as an employer responsibility and relied primarily on institutional supply [17,18]. Therefore, this finding highlights a potential distinction between workers’ perceived responsibility for using protective equipment and their perceived responsibility for their procurement. Although knowledge of ocular hazards and the importance of protective eyewear was high, consistent use of ocular protection was uncommon. Only 7.1% of workers reported consistent eye protection use, despite 97.6% recognizing the need for eye protection. This knowledge–practice gap is consistent with findings from comparable occupational groups in Ghana and sub-Saharan Africa [11,12,16]. The COM-B model of behaviour change provides a useful framework for interpreting this gap. It proposes that behaviour is influenced by three interacting conditions: capability, or the knowledge and skills required to perform a behaviour; opportunity, referring to external factors that enable or constrain the behaviour; and motivation, encompassing the processes that influence an individual’s decision to act [19]. In this study, the high knowledge scores suggest that basic capability was present, but the limited availability of task-appropriate protective eyewear points to an important opportunity-related barrier. Motivation may also have been relevant, given that only 23.5% of workers were willing to personally purchase or contribute toward PPE procurement. Thus, the low uptake of ocular protection may reflect barriers beyond knowledge alone, particularly access to appropriate equipment and perceptions of responsibility for its provision. Furthermore, workers who routinely used general protective equipment were significantly more likely to use ocular protection (Fisher’s exact test, p = 0.011), whereas age, occupation, and educational level were not significantly associated with eyewear use. This finding suggests that ocular protection may be more effectively promoted within a broader workplace safety culture rather than through ocular-safety education alone [3,20].

Ocular injuries were reported by 23.5% of workers within the recall periods assessed, lower than rates reported among welders in Nigeria [21], potentially reflecting differences in task-specific exposure and hazard intensity. Health-seeking behaviour following injury was limited: of the 20 workers who reported an injury, only three sought care at a hospital and two at a pharmacy, while most managed the injury by irrigating the eye with water. While immediate irrigation is an appropriate first-response measure for many ocular exposures, particularly chemical splashes, injuries involving retained foreign bodies or persistent symptoms may require professional assessment. Similar limitations in health-seeking behaviour have been reported in India [22], highlighting the need for targeted training on appropriate first aid and when to seek professional care. Age was not significantly associated with injury occurrence, contrasting with data from the International Labor Organization (ILO) suggesting that younger workers have a higher risk of workplace injuries [23]. This discrepancy may partly reflect the relatively narrow age distribution of the present sample, although differences in occupational exposures and reporting practices may also contribute. Overall, the findings suggest that improving ocular safety should extend beyond injury prevention to include practical training on immediate first aid and appropriate care-seeking following occupational eye injuries. The findings also indicate that improving ocular safety among maintenance workers at KNUST requires more than increasing awareness of ocular hazards. Limited access to task-appropriate PPE, together with gaps in consistent protective practices, highlights the need for stronger institutional support for ocular protection. Reliable PPE provision, section-specific safety training, and monitoring of occupational safety compliance should therefore be prioritized. Future research should evaluate the effectiveness of such interventions in improving protective practices and reducing ocular injuries.

## Data Availability

Data cannot be made publicly available due to ethical restrictions imposed by the Kwame Nkrumah University of Science and Technology Committee on Human Research, Publication and Ethics (CHRPE), which approved this study (Ref: CHRPE/AP/323/21) on the condition that data gathered be used only for the approved purposes, with any other use requiring the Committee's permission. The small, identifiable nature of the study population (85 maintenance workers across seven operational sections at a single named institution) further limits the extent to which data can be shared without risking participant re-identification. Data requests should be directed to the KNUST Committee on Human Research, Publication and Ethics at.

## Acknowledgements

We thank the workers of the Maintenance and Essential Services Organization (MESO), Kwame Nkrumah University of Science and Technology, for their participation and for generously giving their time despite demanding work schedules. We also acknowledge the support of the MESO administration in facilitating access to staff during data collection.

